# Spread of virus during soccer matches

**DOI:** 10.1101/2020.04.26.20080614

**Authors:** Nikolas S. Knudsen, Manuel M. Thomasen, Thomas B. Andersen

## Abstract

In the present the exposure to virus during soccer matches is calculated. Tracking data from 14 elite matches was used. One player in each match was carrying a virus. The exposure score (measured in seconds) was calculated as time spent closer than 1.5m from the infected player or time spent in an exponentially declining zone where the infected player was positioned earlier. The results reveal that, on average, each player was exposed for 87.8s per match.

## Introduction

Following spread of the Covid-19 disease, most countries in the world and the World health Organization (WHO) have emphasized social distancing among the protective measures. This means that sports, in which the athletes are in close contact, are no longer possible. Soccer is one of these sports.

The SARS-CoV-2 virus, as other viruses, is generally believed to spread through contact or indirectly through the air (Weber and Stilianakis 2008). The spread through contact occur when particles from an infected person attach to a surface or an object and other people then touch that surface or object before touching their own eyes, nose or mouth. Furthermore, virus is transmitted through direct contact. The spread through the air occur when an infected person expire particles which are then inspired by other persons. Particles are expired when an infected person coughs, sneezes or through heavy breathing, talking or shouting (Tang 2015). However, it is not clear how far the particles travel through the air or how long they are viable in the air. A general guideline regarding social distancing is that persons should keep a distance of no less than between 1m (WHO) and 2m (National Health Service, United Kingdom).

It is well established that people playing sports are healthier and have a lower risk of numerous diseases (Pedersen and Saltin 2015). Accordingly, it can be argued, that the lack of possibilities for doing sports is detrimental to the public health. Hence, when a virus spreads through the population, health benefits from social distancing should be weighed against the decline in physical activity through sports. However, the probability of virus spread in soccer is not yet established.

Accordingly, in the present study we try to describe a method to estimate the exposure to virus for players during team sports if a player on the field is infected. As a case we try to use this method to determine exposure during soccer matches and identify structural differences in exposure due to player positions.

## Methods

Data was retrieved from one random match at each stadium in the Danish football league (The Danish Superliga) in the 2018/2019-season. Accordingly, data from 14 matches was used. Player position data was collected using a semiautomatic multiple-camera tracking system (Tracab, ChyronHego®). Data was captured at 25 Hz. Player position data was tracked in 2 dimensions (x and y coordinate). Data was filtered using a Butterworth fourth-order low-pass filter with a cut-off frequency of 0.24 Hz filtered the x and y coordinates of the player, using a build in MatLab-function (The MathWorks, inc., New York, USA).

In order to calculate the risk of being infected we calculate a Danger Zone (DZ). We use a distance of 1.5m and calculate the time a player is within this distance from an infected player. Furthermore, we let a tail follow the infected player; a zone where the infected player was positioned time, t, ago. The tail that follows a player is modelling the decline in the amount of virus that stay airborne. Gravity pulls the droplets towards the ground and air resistance opposes this motion. This is modelled as an exponential decline in exposure score and our function is based on the studies by Wells (1984) and Wang et al. (2020).The danger value of this tail is exponentially declining with a half-life of 2s. This means that if a player is positioned within a radius of where an infected player was positioned 2 seconds ago, then the danger value is 0.5 (see figure 1 for an illustration).

**Figure 1.**
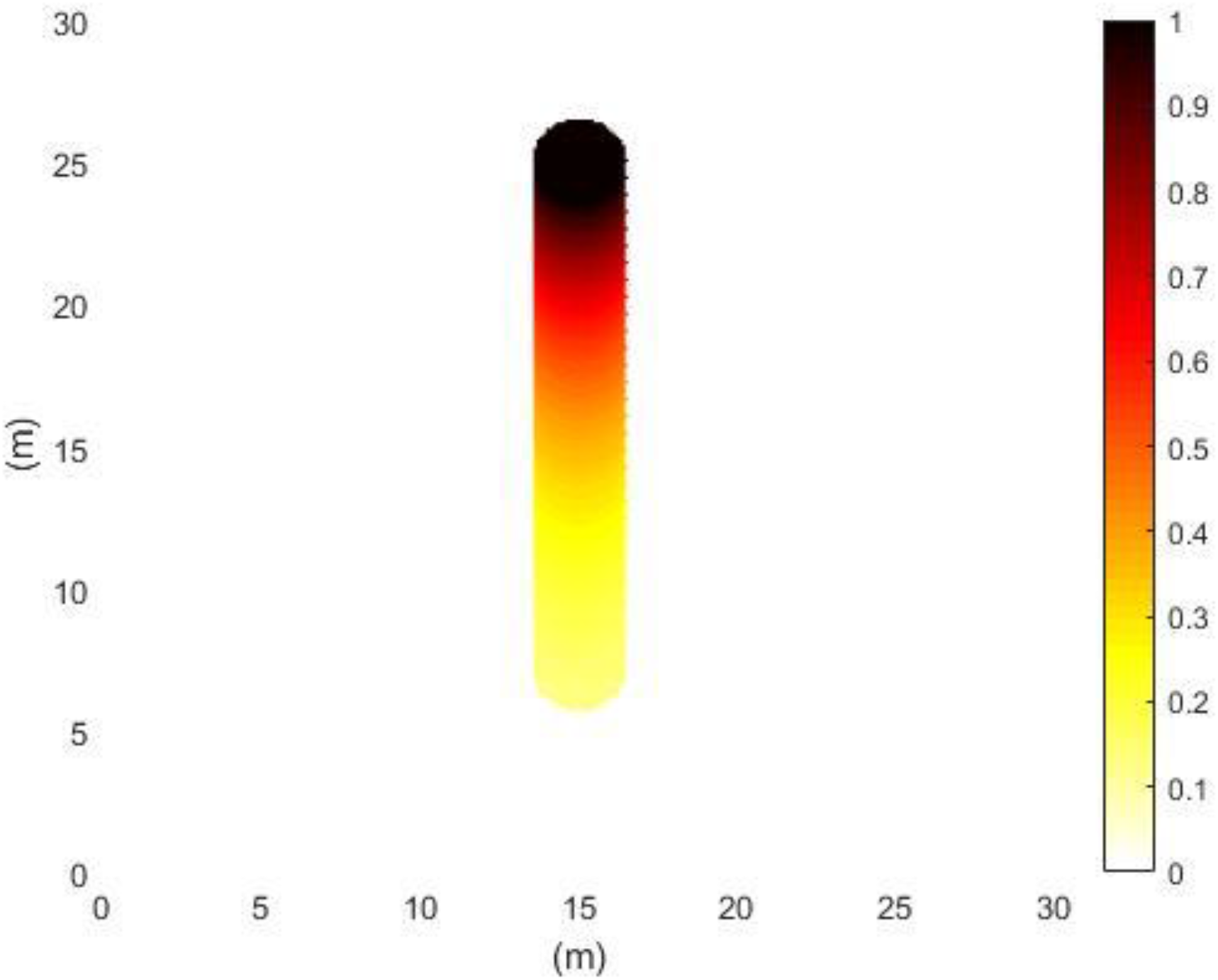
Model of the calculation of the exposure score within one time-frame. The modelled player was running at a constant speed of 3 m s^-1^ in the direction of the y-axis, i.e. the player is positioned at coordinate (15,25). The color-grading shows the exposure score, both close to the player (<1.5m) and in a tail following the player.

For every point in time throughout the match, the DZ and the tail of an infected player is drawn and players who are within one of these zones are given a score; a score of 1 if they are within a distance of 1.5m (DZ-score) and a score of 0.5 if they are within 1.5m of the position an infected player was at 2s ago etc. (tail-score). If a player is within more zones at the same time (ie. stationary player) the score is then determined as the maximal score of the zones. Accordingly, the maximal score at any time and position is 1. An exposure-score is then calculated as the sum of all scores divided by the sample frequency (25Hz). This can be translated as how much time a player spent in a risk zone through a match. For example, a score of 55 corresponds to the player standing within a distance of less than 1.5m from the infected player for 55 seconds.

The calculations were performed with one infected player in each match and repeated until every player had been the infected one. In 14 matches, a total of 15750 exposure-scores were calculated. As these 14 matches were performed by a variety of teams in the Danish Superliga player positions are generalised to the following positions: goalkeeper (GK), defensive backs (LB, CB, and RB), wings and midfielders (LW, MF, and RW), and forwards and strikers (LF, ST, RF). This may result in some teams having multiple players on the same position, as a team playing the 4-4-2 system would have 2 CB’s, 2 MF’s and 2 ST’s, while a team playing the 3-5-2 system would have 2 LW’s and 2 RW’s.

Data from the exposure scores were correlated to the time played and a linear regression expresses the fit, and the exposure-score as a function of time played in one half of a match. Furthermore, when presenting mean and confidence intervals, the exposure score is normalised to the duration of a whole match (90 minutes).

### Statistical analysis

Data is represented as MEAN (95% confidence interval). Exposure scores of positions with different sources of infection are compared using the student T-test with a significance level set at 95%.

## Results

The playing time correlates to the exposure score (p<0.001) (Figure 2). The mean score per 90 minutes (one match) for each player was 87.8s (87.0; 89.6). The highest exposure score was 656.9s and the lowest score was 0s.

**Figure 2.**
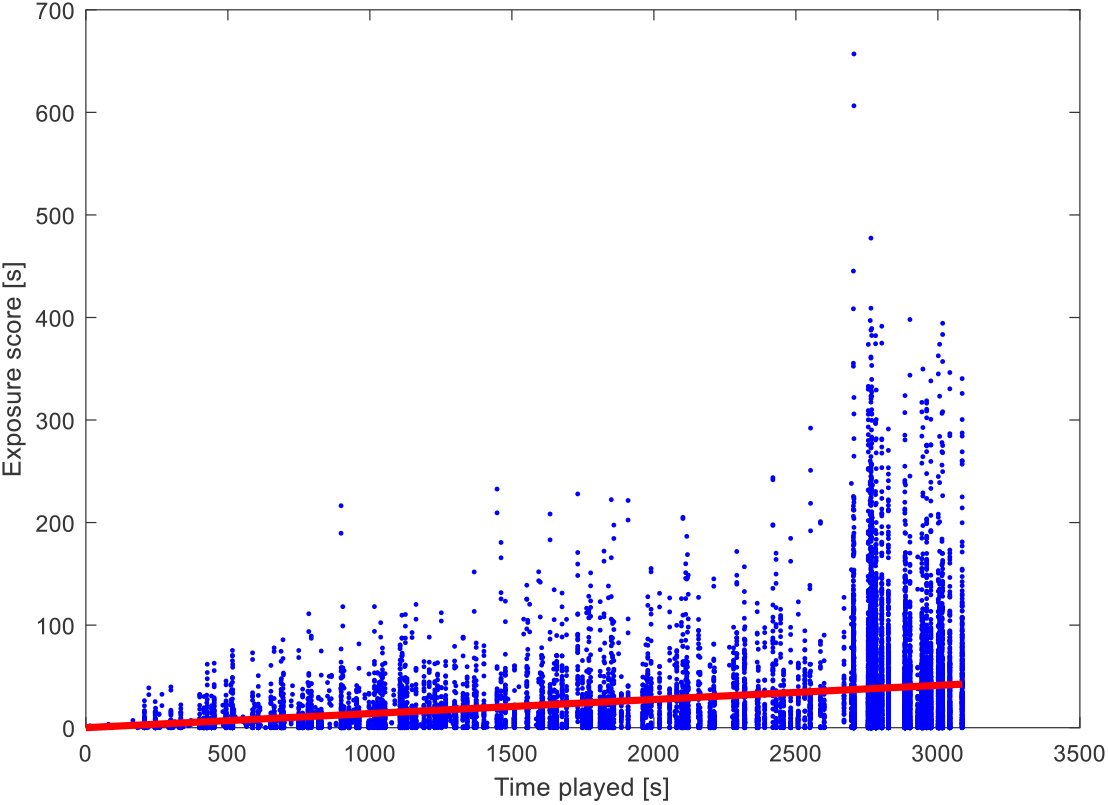
Data from all combinations of players and 1 infected player during one half of a game. The x-axis shows the time played and the y-axis the exposure score.

Exposure score correlates to the position of the infected and non-infected. Mean if player on your own team is infected the mean exposure score was 61.8s (60.4; 63.2) while if a player on opponent team is infected the mean exposure score almost doubles to 111.4 s (108.2; 114.6). The positions resulting in the highest exposure score regardless of which player is infected were Strikers and Center-backs while the Goalkeepers had significantly lower exposure score than any other position (P<0.05) (figure 3). Exposure score of any position when accounting for which player (position and team) is infected can be seen in figures 4 A (infection source on own team) and 4 B (infection source on opponent team). All results can be seen in table 1.

**Figure 3.**
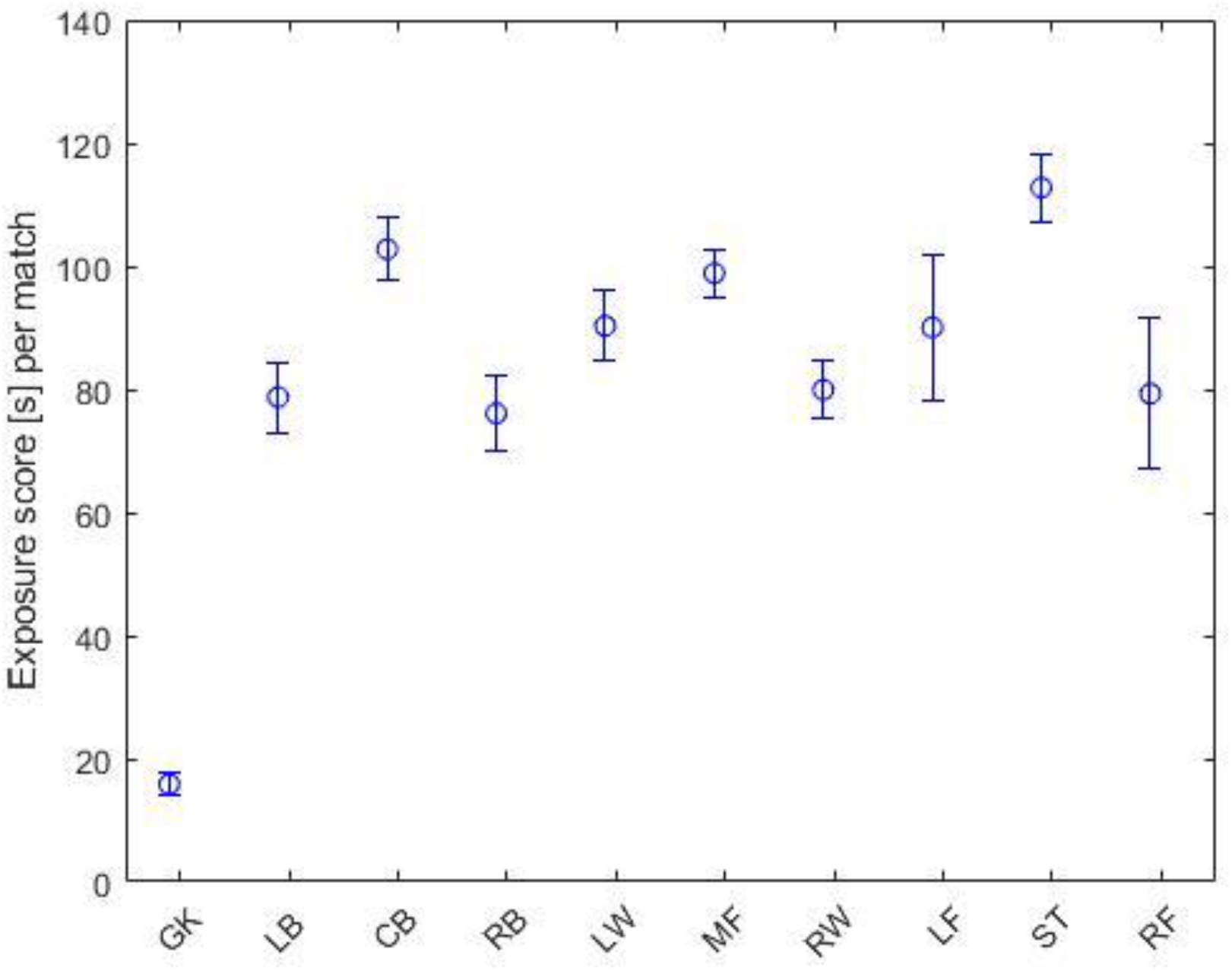
Exposure score of players regardless of the source of infection. The x-axis shows the positions and the y-axis the exposure score.

**Figure 4 A.**
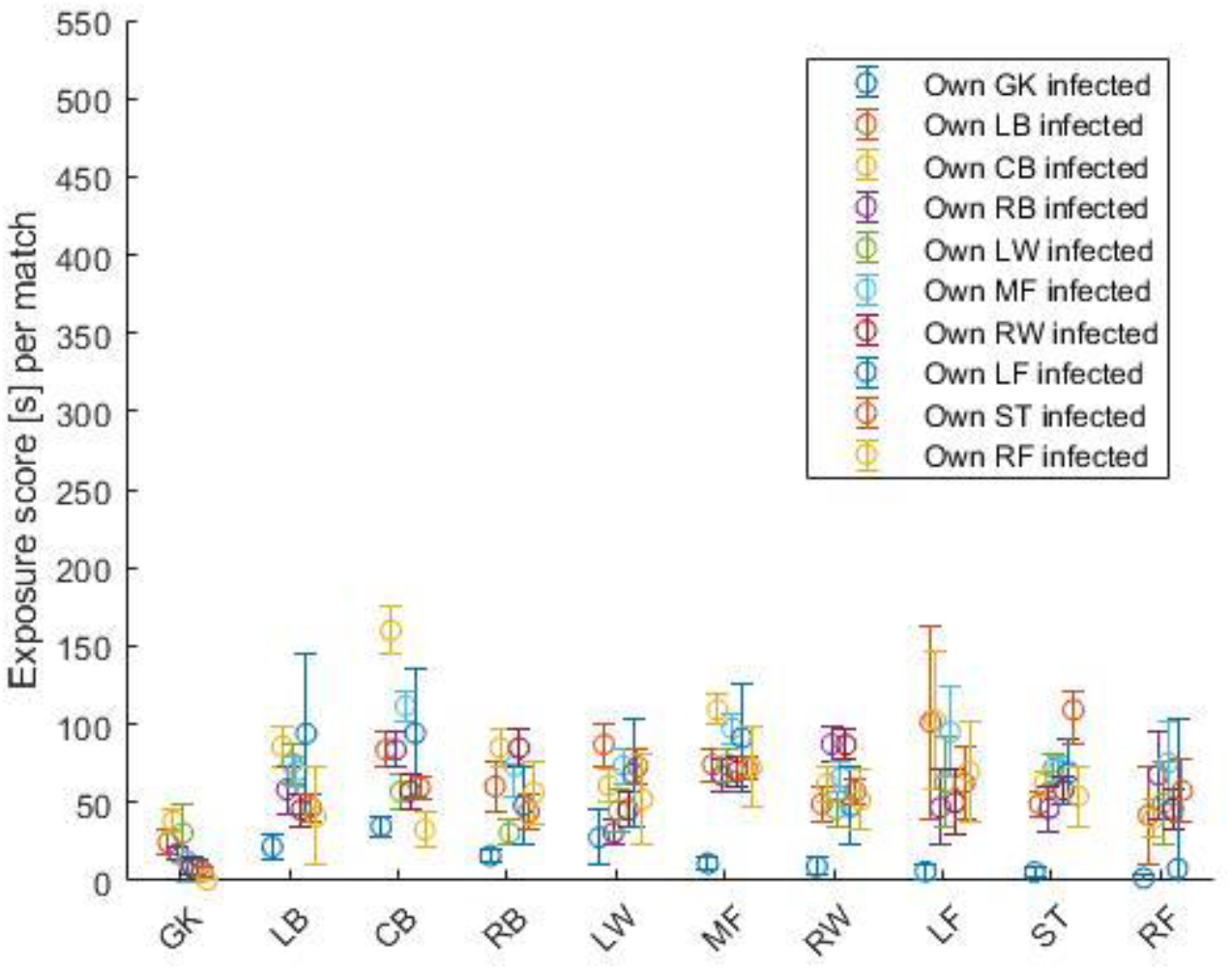
Exposure score of the individual positions for any other player on own team infected. The x-axis shows the positions and the y-axis the exposure score.

## Discussion

Our results show that, on average, a player is positioned within an exposure zone for 1 minute and 28 seconds (87.8s) during a soccer match. We have not been able to find any data on the minimum exposure time before infection. Furthermore, the exposure time corresponds linearly to the playing time. This means that if matches are shorter, then the exposure will be smaller.

Both results can be used in the ongoing discussion on the potential re-opening of sports facilities and the playing of soccer matches. Our analysis does not include transmission of virus through contact. This phenomenon will mainly occur when performing throw-ins or during tackles. On the other hand, the analysis does include the celebrations after scoring. Players from the scoring team usually get in close contact when celebrating a goal. In the analysed season an average of 2.6 goals were scored per match. The exposure score can be smaller if players keep their social distance when the ball is not in play.

Our results further show a disparity in exposure score depending on which team and position the infected player plays. A lower exposure score when the infected players is on ones own team suggests tactical structures causing larger mean distance between players on a team. As defences tend to focus on covering larger areas and offences tend to space out in order to stretch the opponent’s defence these structures causes what could be referred to as tactical caused social distancing, causing the exposure score to decrease. In contrast, exposure scores rise when the infected player is on the opponent team. This might arise due to players wanting to cover the opponent team, causing the distance between players to decrease. This is further supported by figure 4 B showing increased exposure score when a players’ direct opponent is infected (e.g. increased exposure score of CB’s when opponent ST is infected).

**Figure 4 B.**
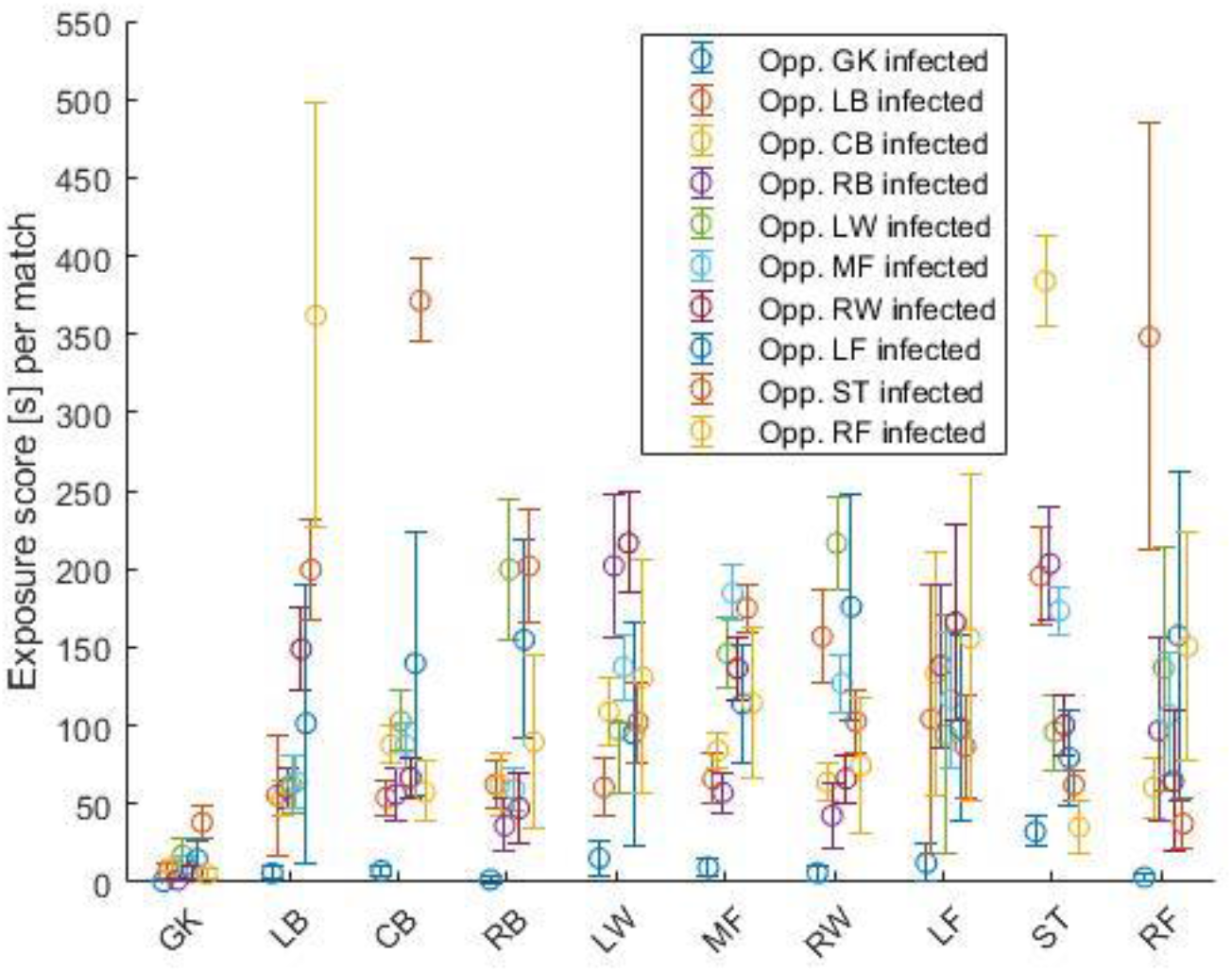
Exposure score of the individual positions for any other player on opponent team infected. The x-axis shows the positions and the y-axis the exposure score

Our analysis only include one infected player. If more players are infected the results can simply be added together from the exposure scores of the infected players.

In the present study we used data from elite soccer matches. It is obvious that players at a different level or in different age groups do not move as elite players. Accordingly, it is uncertain how the exposure scores will be for other groups.

Based on our analyses, we are not able to conclude whether or not the soccer players a in a high risk of being infected during matches. Our calculated exposure time corresponds to standing within 1.5m of an infected person for less than 1½ minute.

## Data Availability

Data was shared for research purposes by the Danish League (Divisionsforeningen) who collect tracking data fra all matches.

## Acknowledgements

The authors would like to thank the Danish League (Divisionsforeningen) for letting us access tracking data.

