## Supplementary material for "Spread of virus during soccer matches": Table 1

| | GK | LB | CB | RB | LW | MF | RW | LF | ST | RF | | Any position |
|---|---|---|---|---|---|---|---|---|---|---|---|---|
| Regardless of infected | 15.9 (13.9; 17.8) | 78.7 (73.0; 84.4) | 102.8 (97.6; 107.9) | 76.1 (70.1; 82.1) | 90.3 (84.6; 96.0) | 98.9 (95.0; 102.8) | 79.9 (75.2; 84.7) | 90.1 (78.2; 101.9) | 112.8 (107.2; 118) | 79.3 (67.1; 91.4) | | 87.8 (86.0; 89.6) |
| Any own infected | 18.3 (15.4; 21.2) | 60.7 (56.3; 65.1) | 79.2 (75.1; 83.3) | 57.0 (52.6; 61.4) | 60.5 (55.9; 65.1) | 75.5 (72.0; 79.0) | 55.7 (51.7; 59.7) | 68.1 (56.3; 79.9) | 61.1 (57.7; 64.5) | 50.9 (43.0; 58.98) | | 61.6 (60.4; 63.2) |
| Any opponent infected | 13.6 (11.0; 16.2) | 95.2 (85.2; 105.2) | 124.4 (115.3; 134) | 93.6 (83.0; 104.2) | 117.1 (107.4; 127) | 120.1 (113.6; 127) | 101.6 (93.6; 109.6) | 109.7 (90.2; 129.2) | 159.5 (150.0; 169) | 104.7 (83.4; 126.0) | | 111.4 (108.2; 115) |
| Own GK infected | - | 21.6 (14.1; 29.1) | 34.7 (28.1; 41.3) | 16.0 (11.4; 20.5) | 27.8 (10.3; 45.3) | 11.1 (7.6; 14.5) | 9.1 (3.6; 14.7) | 6.0 (1.1; 10.8) | 5.6 (2.6; 8.7) | 1.7 (-0.7; 4) | | 16.6 (13.9; 19.3) |
| Own LB infected | 24.5 (16.2; 32.8) | - | 83.8 (72.2; 95.4) | 60.4 (44.5; 76.2) | 87.1 (73.3; 100.8) | 74.3 (63.8; 84.9) | 49.1 (37.2; 60.9) | 101.3 (40.0; 162.6) | 49.1 (40.8; 57.5) | 42.0 (10.7; 73.3) | | 63.9 (59.3; 68.5) |
| Own CB infected | 38.1 (31.0; 45.1) | 86.0 (73.7; 98.2) | 159.9 (144.8; 175) | 85.2 (73.4; 97.0) | 61.0 (50.3; 71.7) | 109.3 (99.6; 119.0) | 61.8 (50.6; 73.0) | 102.7 (59.1; 146.3) | 62.3 (54.4; 70.1) | 39.8 (28.2; 51.4) | | 81.3 (77.2; 85.4) |
| Own RB infected | 17.5 (12.9; 22.0) | 58.3 (43.1; 73.6) | 83.6 (72.6; 94.7) | - | 30.8 (22.8; 38.7) | 67.2 (57.3; 77.2) | 87.3 (75.9; 98.7) | 46.9 (23.0; 70.9) | 46.2 (31.8; 60.6) | 67.8 (39.5; 96.1) | | 58.4 (53.7; 63.1) |
| Own LW infected | 30.5 (11.3; 49.7) | 75.1 (62.5; 87.8) | 56.9 (46.3; 67.6) | 30.7 (22.5; 38.8) | 45.0 (31.6; 58.3) | 74.2 (64.0; 84.3) | 45.9 (34.2; 57.6) | 63.0 (34.4; 91.7) | 72.0 (62.4; 81.5) | 49.7 (23.2; 76.3) | | 58.7 (54.3; 63.1) |
| Own MF infected | 12.5 (9.0; 16.0) | 70.2 (60.5; 79.9) | 111.7 (102.2; 121.2) | 63.2 (53.8; 72.6) | 73.6 (62.5; 84.7) | 97.6 (88.0; 107.2) | 66.1 (56.5; 75.6) | 95.1 (66.0; 124.1) | 71.4 (63.8; 79.1) | 75.3 (48.3; 102.3) | | 74.8 (71.3; 78.3) |
| Own RW infected | 9.3 (4.0; 14.5) | 45.5 (35.1; 55.9) | 57.2 (46.4; 68.0) | 84.9 (73.4; 96.4) | 45.3 (33.8; 56.9) | 70.1 (60.8; 79.4) | 86.9 (77.3; 96.5) | 50.0 (29.1; 71.0) | 58.0 (49.6; 66.4) | 46.0 (32.9; 59.2) | | 55.2 (52.3; 59.1) |
| Own LF infected | 8.0 (3.0; 13.1) | 94.1 (43.0; 145.1) | 94.3 (53.1; 135.4) | 48.2 (23.0; 73.3) | 68.7 (34.8; 102.7) | 91.4 (56.6; 126.2) | 47.6 (22.9; 72.3) | - | 69.1 (48.5; 89.8) | 71.7 (39.7; 103.6) | | 67.2 (55.8; 78.6) |
| Own ST infected | 6.1 (3.1; 9.1) | 46.9 (38.2; 55.6) | 59.6 (52.1; 67.1) | 44.1 (33.2; 54.9) | 73.1 (62.0; 84.3) | 71.8 (64.2; 79.3) | 57.0 (48.9; 65.1) | 62.6 (39.4; 85.8) | 109.2 (97.2; 121.1) | 57.7 (37.9; 77.5) | | 60.4 (57.0; 63.8) |
| Own RF infected | 0.5 (-0.1; 1.1) | 41.0 (9.7; 72.4) | 32.7 (22.0; 43.3) | 56.5 (36.3; 76.8) | 52.0 (23.4; 80.7) | 72.3 (46.6; 98.1) | 51.6 (32.5; 70.7) | 69.6 (38.1; 101.1) | 54.0 (34.6; 73.5) | - | | 48.5 (40.7; 56.3) |
| Opponent GK infected | 0.6 (-0.1; 1.4) | 5.9 (1.9; 9.8) | 7.3 (4.7; 10.0) | 1.7 (0.0; 3.3) | 15.4 (4.2; 26.6) | 9.5 (4.2; 14.9) | 6.1 (1.6; 10.6) | 12.3 (0.1; 24.5) | 32.4 (23.1; 41.7) | 3.2 (0.9; 5.4) | | 11.6 (9.2; 14.0) |
| Opponent LB infected | 6.9 (2.0; 11.8) | 55.6 (17.0; 94.2) | 54.0 (42.4; 65.6) | 62.4 (47.5; 77.3) | 60.6 (42.6; 78.6) | 66.0 (50.1; 81.9) | 156.8 (127.1; 187) | 104.3 (17.8; 190.8) | 195.5 (163.7; 227) | 348.4 (211.9; 485) | | 95.6 (85.6; 105.6) |
| Opponent CB infected | 10.2 (7.1; 13.2) | 53.2 (42.1; 64.4) | 87.7 (75.5; 99.9) | 62.4 (43.0; 81.9) | 109.1 (88.1; 130.2) | 84.3 (72.3; 96.2) | 64.0 (51.8; 76.1) | 133.5 (55.5; 211.4) | 383.8 (355.4; 412) | 60.5 (41.6; 79.5) | | 126.9 (117.5; 136) |
| Opponent RB infected | 1.5 (0.3; 2.7) | 60.3 (47.1; 73.6) | 56.2 (39.7; 72.7) | 36.2 (19.3; 53.1) | 202.1 (156.3; 248) | 57.3 (44.3; 70.3) | 42.6 (22.1; 63.1) | 138.2 (85.6; 190.8) | 203.5 (167.4; 240) | 96.9 (37.0; 156.7) | | 91.9 (81.5; 102.3) |
| Opponent LW infected | 17.1 (5.4; 28.8) | 62.4 (44.0; 80.8) | 102.9 (83.8; 122.0) | 199.8 (155.4; 244) | 79.2 (56.8; 101.6) | 146.2 (124.0; 169) | 216.5 (186.4; 247) | 94.3 (18.6; 170.0) | 96.1 (72.1; 120.2) | 136.8 (58.8; 214.8) | | 116.9 (107.4; 126) |
| Opponent MF infected | 11.1 (5.6; 16.6) | 64.3 (47.9; 80.7) | 87.5 (73.7; 101.3) | 59.3 (45.7; 73.0) | 137.6 (116.6; 159) | 185.0 (166.9; 203) | 127.1 (109.0; 145) | 116.8 (73.3; 160.3) | 173.5 (158.2; 189) | 106.8 (67.6; 146.0) | | 118.5 (112.0; 125) |
| Opponent RW infected | 6.6 (2.2; 11.0) | 148.9 (122.6; 175) | 66.9 (54.5; 79.3) | 47.3 (24.2; 70.4) | 216.7 (184.5; 249) | 136.5 (116.7; 156) | 66.2 (51.3; 81.1) | 166.1 (103.7; 229) | 100.4 (81.0; 119.8) | 64.5 (19.6; 109.4) | | 102.7 (94.6; 110.8) |
| Opponent LF infected | 14.7 (2.4; 27.0) | 101.6 (12.7; 190.6) | 139.9 (56.1; 223.8) | 154.9 (91.7; 218.2) | 94.8 (23.9; 165.8) | 113.9 (76.5; 151.2) | 175.9 (103.8; 248) | 98.6 (38.5; 158.7) | 79.6 (49.5; 109.6) | 157.6 (52.3; 262.9) | | 111.3 (91.5; 131.1) |
| Opponent ST infected | 38.2 (28.1; 48.4) | 199.7 (168.2; 231) | 371.3 (344.8; 398) | 202.0 (165.3; 239) | 102.4 (76.7; 128.2) | 175.0 (159.4; 191) | 102.6 (82.2; 123.0) | 86.7 (53.8; 119.6) | 62.1 (52.4; 71.9) | 37.5 (20.9; 54.1) | | 159.6 (150.3; 169) |
| Opponent RF infected | 5.3 (2.2; 8.4) | 361.9 (226.1; 497.7) | 57.8 (38.5; 77.1) | 89.8 (34.1; 145.5) | 130.8 (56.2; 205.3) | 114.5 (66.2; 162.8) | 74.8 (31.1; 118.6) | 156.0 (51.6; 260.3) | 35.3 (18.6; 51.9) | 150.5 (77.5; 223.5) | | 106.5 (84.5; 128.5) |

Table 1: Exposure scores from each individual position. Results are presented as mean with 95% confidence intervals in parenthesis.
